# Ocular Manifestations in a Cohort of 43 Patients with KBG Syndrome

**DOI:** 10.1101/2023.05.31.23290743

**Authors:** Drake C. Carter, Ola Kierzkowska, Kathleen Sarino, Lily Guo, Elaine Marchi, Gholson J Lyon

## Abstract

Ophthalmological conditions are underreported in patients with KBG syndrome, which is classically described as presenting with dental, developmental, intellectual, skeletal, and craniofacial abnormalities. This study analyzed the prevalence of four ophthalmological conditions (strabismus, astigmatism, myopia, hyperopia) in 43 patients with KBG syndrome carrying pathogenic variants in *ANKRD11* or deletions in 16q24.3 and compared it to the literature. 43 patients were recruited via self-referral or a private Facebook group hosted by the KBG Foundation. Virtual interviews were conducted to collect a comprehensive medical history verified by medical records. From these records, data analysis was performed to calculate the prevalence of ophthalmological conditions. Strabismus was reported in 10 (23.3%) participants, while astigmatism, myopia, and hyperopia were reported in 12 (27.9%), 7 (16.3%), and 9 (20.9%) participants, respectively. Other reported conditions include anisometropia, amblyopia, and nystagmus. When compared to the literature, the prevalence of strabismus and refractive errors is higher than other studies. However, more research is needed to determine if mutations in *ANKRD11* play a role in abnormal development of the visual system. In patients with established KBG syndrome, screening for misalignment or refractive errors should be done, as interventions in patients with these conditions can improve functioning and quality of life.

## 1 INTRODUCTION

KBG syndrome was first reported in 1975 by Herrman et al. and is named after the surnames of the first three families diagnosed (K, B, and G) [1]. It is associated with dental, developmental, skeletal, and craniofacial abnormalities [2]. Clinical findings in affected individuals include macrodontia, intellectual disability (ID), short stature, brachydactyly, and costovertebral anomalies [3–5]. Other research has expanded the phenotype of KBG syndrome to include the occurrence of congenital heart defects, seizures, and gastrointestinal problems [6–8]. Currently, the prevalence of KBG syndrome is unknown but it is thought to be underdiagnosed due to the nonspecific and wide spectrum of symptoms [4]. Genetic or genomic testing are ways to confirm the diagnosis of suspected KBG syndrome [9].

The genetic etiology of KBG Syndrome is from variants in Ankyrin Repeat Domain (*ANKRD11*) and deletions in 16q24.3 [4]. Variants in *ANKRD11* have also been found in patients clinically diagnosed as possibly having Cornelia de Lange syndrome (CdLS), another developmental disorder that overlaps clinically with KBG syndrome [10].

Among the clinical symptoms commonly attributed to KBG syndrome, ophthalmologic findings are sparsely mentioned in the literature. Refractive errors (myopia, hyperopia, astigmatism) and strabismus have recently been reported in patients [9,11]. Even though ophthalmologic evaluation is recommended in patients after initial diagnosis of KBG syndrome [9], there is a lack of research that investigates the link between KBG syndrome and ophthalmologic symptoms, if one is present. The prompt diagnosis and management of strabismus is essential in preventing permanent amblyopia [12]. Refractive errors should also be promptly addressed to improve visual deficits and life outcomes in pediatric patients [13].

Since KBG syndrome is a rare condition, there have been efforts to sufficiently describe the phenotypical spectrum of diagnosed patients as well as describe novel clinical presentations [14–16]. The aim of this study is to describe the ophthalmological manifestations in a cohort of 43 patients with diagnosed KBG syndrome and compare descriptively the prevalence of refractive errors and strabismus in our cohort to the literature.

## 2 METHODS

43 patients (19 females, 24 males) with a pathogenic variant in *ANKRD11* (n = 41) or deletions in 16q24.3 (n = 2) were interviewed via secure video conference platforms between January 2021 and May 2022. Video interviews were conducted to collect a thorough medical history of each patient, including problems with vision or other ophthalmological complaints. Past medical records, including genetic testing results confirming variants in *ANKRD11*, were collected as well. Interviews were approximately one to two hours long and consisted of questions investigating the function of all organ systems as well as a characterization of their social, cognitive, and developmental history. All interviewed research participants were self-referred or recruited via a private Facebook group monitored and hosted by the KBG foundation. Written consent was obtained for the collection of medical records.

Microsoft Excel was used to organize and determine prevalence data of ophthalmological conditions reported by interviewed participants. Of most relevance was the prevalence of strabismus, astigmatism, myopia (near-sightedness), and hyperopia (far-sightedness). Other conditions were reported by our cohort, including anisometropia (n=3), nystagmus (n=1), amblyopia (n=2), and cortical visual impairment (n=1), but were not deemed relevant due to a low number of affected individuals in our cohort.

A PubMed search was conducted to find prevalence data of strabismus, astigmatism, myopia, and hyperopia of populations with similar demographics to ours (pediatric and non-Hispanic). Initial search criteria included the keywords “KBG” AND “astigmatism OR hyperopia OR myopia OR strabismus” to gather literature regarding the known reports of visual impairment in KBG Syndrome. A second search was conducted using the keywords “astigmatism OR hyperopia OR myopia OR strabismus” AND “prevalence” to collect and analyze studies describing the prevalence of these visual problems in various patient populations. Studies were prioritized if they analyzed populations that were Caucasian, non-Hispanic, and under 18, as this matches the demographics of the majority of our cohort (mean age 13.4, 95.3% Caucasian, 86.0% non-Hispanic). However, studies utilizing patient populations outside these parameters were also considered for comparison to their appropriate demographic in our cohort. Prevalence data from the literature for each condition was compared to our calculated prevalence, and a descriptive statistical analysis was performed.

## 3 RESULTS

### 3.1 Participant demographics

Patients come from 12 countries (55.8% from The United States) with a mean age of

13.4 years (range 1-59). Amongst our cohort, 41 patients (95.3%) were Caucasian, 37 patients (86.0%) were non-Hispanic, and 32 patients (74.4%) were under the age of 18. **Table 1** outlines the demographic information of interviewed participants with KBG syndrome.

**Table 1.**
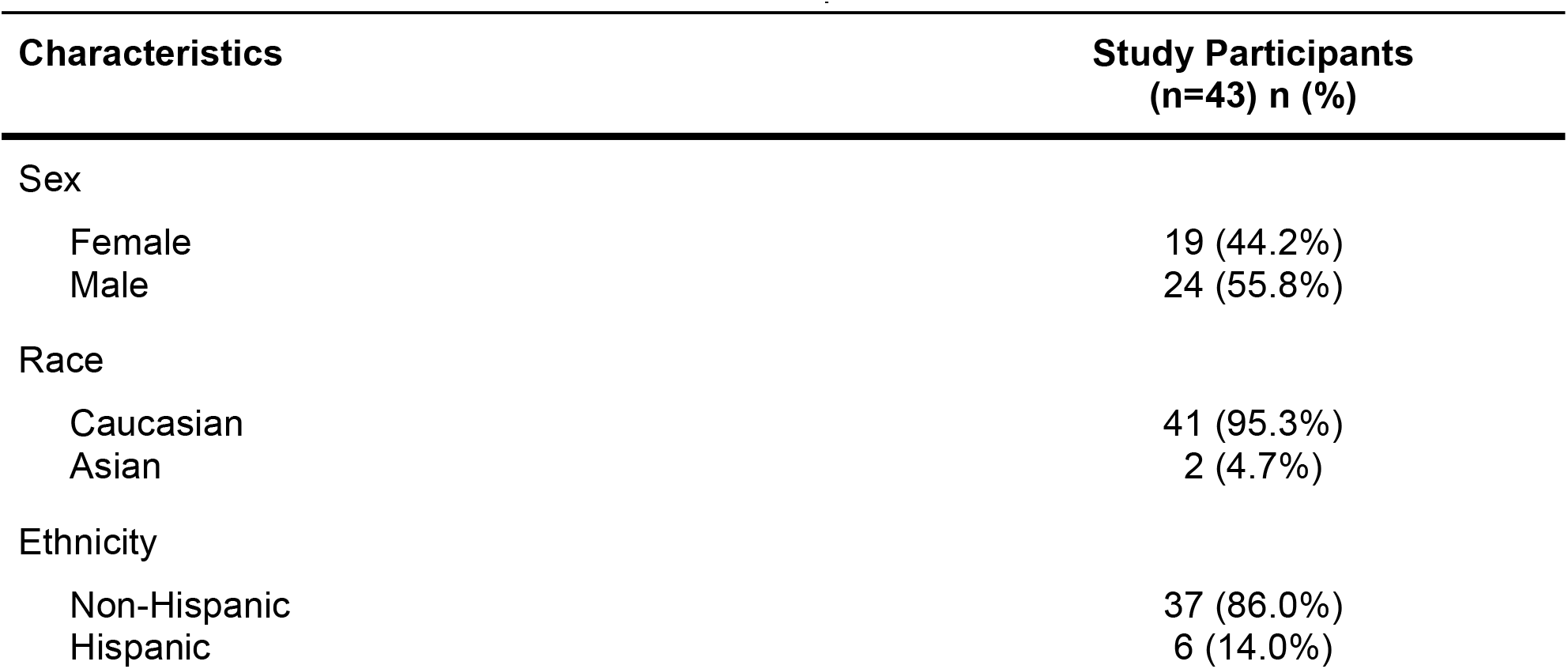
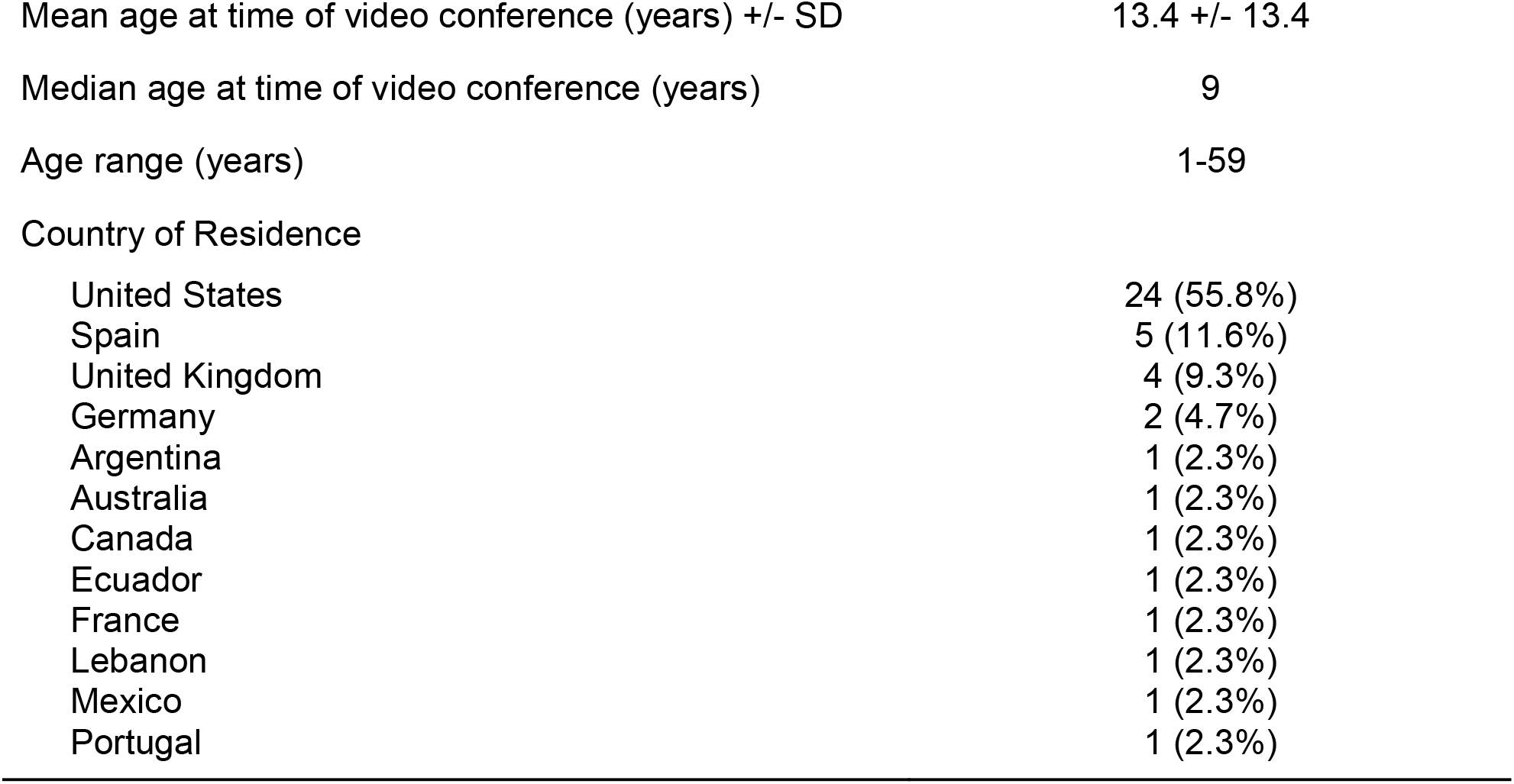
Demographic information of interviewed patients.

| Characteristics | Study Participants<br>(n=43) n (%) |
| --- | --- |
| Sex |  |
| Female | 19 (44.2%) |
| Male | 24 (55.8%) |
| Race |  |
| Caucasian | 41 (95.3%) |
| Asian | 2 (4.7%) |
| Ethnicity |  |
| Non-Hispanic | 37 (86.0%) |
| Hispanic | 6 (14.0%) |
| Mean age at time of video conference (years) +/- SD | 13.4 +/- 13.4 |
| Median age at time of video conference (years) | 9 |
| Age range (years) | 1-59 |
| Country of Residence |  |
| United States | 24 (55.8%) |
| Spain | 5 (11.6%) |
| United Kingdom | 4 (9.3%) |
| Germany | 2 (4.7%) |
| Argentina | 1 (2.3%) |
| Australia | 1 (2.3%) |
| Canada | 1 (2.3%) |
| Ecuador | 1 (2.3%) |
| France | 1 (2.3%) |
| Lebanon | 1 (2.3%) |
| Mexico | 1 (2.3%) |
| Portugal | 1 (2.3%) |

### 3.2 Variants in *ANKRD11*

All patients were confirmed to have variants in *ANKRD11* or deletions of 16q24.3, which is criteria for a diagnosis of KBG syndrome. **Table 2** outlines the genetic variants of each proband, as well as their assigned proband number. As of October 2022, proband 24 is awaiting genetic testing to confirm the cDNA change, protein change, and inheritance pattern of their variant in *ANKRD11*.

**Table 2.** *ANKRD11* Variants of Participants

| Proband | Age | Sex | cDNA Change | Protein Change | Inheritance |
| --- | --- | --- | --- | --- | --- |
| 1 | 0-4 | M | c.1903_1907del | p.(Lys635fs) | de novo |
| 2 | 5-9 | M | c.3770_3771delAA | p.(Lys1257RArgfs*25) | unknown |
| 3 | 0-4 | F | c.1181delA | p.(Asn394Ilefs*33) | de novo |
| 4 | 5-9 | F | c.2329_2332del | p.(Glu777Argfs*5) | unknown |
| 5 | 15-19 | M | c.6472G>T | p.(E2158*) | de novo |
| 6 | 10-14 | M | c.7354C>G | p.(Arg2452Gly) | de novo |
| 7 | 30-34 | F | c.5233_5234del | p.(Ser1745Hisfs*51) | maternal |
| 8 | 60-64 | F | c.5233_5234del | p.(Ser1745Hisfs*51) | unknown |
| 9 | 5-9 | F | c.2404_2407del | p.(Leu802Lysfs*60) | de novo |
| 10 | 21-24 | M | c.4384dupA | p.(Arg1462Lysfs*92) | unknown |
| 11 | 5-9 | M | 158 kB loss in 16q24.3 | - | unknown |
| 12 | 5-9 | M | c.6628G>T | p.(Glu2210*) | unknown |
| 13 | 0-4 | F | c.3991del | p.(Asp1331Thrfs*14) | de novo |
| 14 | 20-24 | F | c.2177_2178del | p.(Lys0726Argfs*15) | de novo |
| 15 | 20-24 | M | c.1018dup | p.(Thr340Asnfs*9) | unknown |
| 16 | 5-9 | F | c.1903_1907del | p.(Lys0635Glnfs*26) | de novo |
| 17 | 10-14 | M | c.7825C>T | p.(Gln2609*) | de novo |
| 18 | 5-9 | M | c.4489_4490del | p.(Arg1497Glyfs*56) | de novo |
| 19 | 10-14 | M | c.6015dupA | p.(Gly2006Argfs*26) | de novo |
| 20 | 0-4 | F | c.1318C>T | p.(Arg440*) | de novo |
| 21 | 5-9 | F | c.2409_2412 del | p.(Glu0805Argfs*57) | unknown |
| 22 | 5-9 | M | c.3770_3771delAA | p.(Lys1257Argfs*25) | de novo |
| 23 | 5-9 | M | c.7607G>A | p.(Arg2536Gln) | de novo |
| 24 | 5-9 | F | Not reported | Not reported | de novo |
| 25 | 35-39 | F | c.1756G>A | p.(Val0586Met) | unknown |
| 26 | 0-4 | M | c.1756G>A | p.(Val0586Met) | maternal |
| 27 | 0-4 | M | c.1756G>A | p.(Val0586Met) | maternal |
| 28 | 5-9 | M | c.1318C>T | p.(Arg0440*) | de novo |
| 29 | 0-4 | M | c.2175_2178delCAAA | p.(Asn0725Lysfs*23) | de novo |
| 30 | 5-9 | F | c.2398_2401delGAAA | p.(Glu800Asnfs*62) | de novo |
| 31 | 15-19 | M | c.7789A>T | p.(Lys2597*) | paternal |
| 32 | 55-59 | M | c.7789A>T | p.(Lys2597*) | unknown |
| 33 | 10-14 | F | c.2273dup | p.(Arg759Glu*23fs) | unknown |
| 34 | 20-24 | F | c.6596_6597insA | p.(Ala2201Cysfs*6) | de novo |
| 35 | 10-14 | F | c.5659C>T | p.(Gln1887*) | unknown |
| 36 | 5-9 | M | c.5227C>T | p.(Gln1743*) | maternal |
| 37 | 15-19 | M | c.3224_3227delAAAG | p.(Glu1075Glyfs*242) | unknown |
| 38 | 5-9 | F | c.1977C>G | p.(Tyr0659*) | unknown |
| 39 | 5-9 | M | c.2409_2412del | p.(Glu805Argfs*57) | de novo |
| 40 | 20-24 | M | c.3770_3771del | p.(Lys1257Argfs*25) | de novo |
| 41 | 0-4 | M | c.4396_4397 | p.(Arg1466Glyfs*87) | de novo |
| 42 | 5-9 | F | c.1893delA | p.(Lys0631fs) | de novo |
| 43 | 30-34 | F | c.5227C>T | p.(Gln1743*) | unknown |

### 3.3 Strabismus, Astigmatism, Myopia, and Hyperopia

The prevalence of strabismus in our entire cohort was calculated to be 23.3% (n=10). In non-Hispanic participants (n=37), the prevalence of strabismus was 18.9% (n=7). For our participants who were under 18 (n=32), the prevalence of strabismus was 28.1% (n=9).

Among our entire cohort, the prevalence of astigmatism was 27.9% (n=12). The prevalence of astigmatism in our non-Hispanic patients (n=37) was 24.3% (n=9). For our participants who were under 18 (n=32), the prevalence of astigmatism was 31.3% (n=10).

For myopia (nearsightedness), the prevalence among our entire cohort was 16.3% (n=7), while the prevalence among our non-Hispanic (n=37) participants was 18.9% (n=7). The prevalence of myopia among patients under 18 (n=32) was 21.88% (n=7).

The prevalence of hyperopia (farsightedness) in our entire cohort was 20.9% (n=9).

Among those of non-Hispanic origin (n=37), the prevalence was 16.2% (n=6). The prevalence of hyperopia among patients under 18 (n=32) was 21.9% (n=7).

Figure 1 outlines the results of each of these four ophthalmological conditions. Several of our patients also reported having more than one of the described eye conditions (strabismus, astigmatism, myopia, or hyperopia), with 25.6% (n=11) reporting the presence of at least two conditions and 7.0% (n=3) reporting at least 3 conditions.

**Figure 1.**
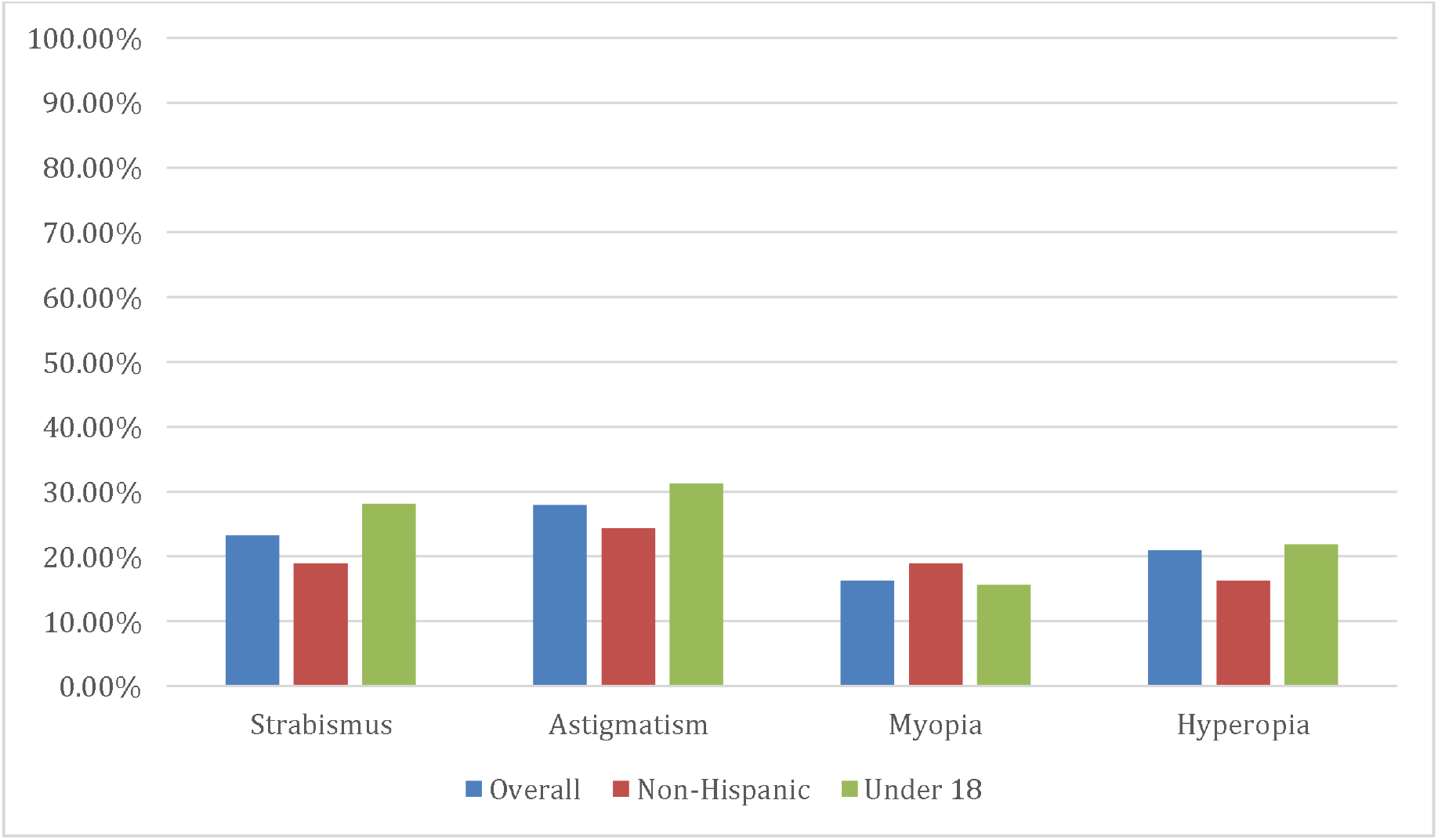
Percentage prevalence of strabismus, astigmatism, myopia, and hyperopia among the entire cohort, as well as non-Hispanic and pediatric (<18) patients.

Other eye conditions were reported among our cohort, but to a lesser extent. These conditions included anisometropia (n=3), nystagmus (n=1), unspecified blurred vision (n=2), amblyopia (n=2), ptosis (n=1), Salzmann’s nodular degeneration (n=1), intermediate uveitis (n=1), cortical visual impairment (n=1), ocular immaturity (n=1), and difficulty adjusting from light to dark (n=1). 62.8% (n=27) of all participants reported having at least one ophthalmological condition.

## 4 DISCUSSION

Although ophthalmological findings have been reported in some cases of KBG syndrome [2,11,17,18], there is currently a lack of research that estimates the incidence of visual problems among those with diagnosed KBG syndrome, or determines if there is a link between KBG syndrome and visual impairments. Our analysis of visual impairments in patients with KBG syndrome hints that a significant relationship may exist between the two.

Among our entire cohort, the prevalence of strabismus (23.3%) is much higher than studied pediatric populations, which have shown to have a prevalence between 2.1 and 3.6% depending on race and ethnicity [19–21]. This prevalence is also much higher than reported by an analysis from the Intelligent Research in Sight (IRIS) Registry, which determined that 2.75% of over 30 million patients in the United States had a diagnosis of strabismus [22].

When comparing our cohorts’ prevalence of astigmatism (27.9%), it proved to also be higher than several studies of other populations. The prevalence range, like strabismus, varied with race and ethnicity between 6.3% in non-Hispanic white (NHW) children and 16.8% in Hispanic children in studies done by the Multi-Ethnic Pediatric Eye Disease Study Group [23,24]. In comparison, 15.6% of our non-Hispanic patients and 31.3% of our Hispanic patients reported a diagnosis of astigmatism. Another study determined the prevalence of astigmatism of nearly 19,000 school-aged children in Philadelphia to be 7.8%, which is much lower than our cohort [25].

The previous studies also determined that the prevalence of myopia (nearsightedness) of various populations to be 1.20% in NHW children, 3.70% in Hispanic children, and 9.4% in Philadelphia school-aged children [23,25,26]. Our overall cohort and our non-Hispanic participants clearly had much higher rates of 16.28% and 21.88%, respectively. Interestingly, none of our Hispanic patients reported myopia.

For studies done on the prevalence of hyperopia amongst populations, the data was quite variable. Studies have reported a range 2.4 to 25.65% among children of various races and ethnicities [23,25]. Another study showed that the prevalence of hyperopia among a cohort of school-aged children ranged from 12.7 to 19.3% among Hispanic and white children, respectively [27]. Our cohort also showed wide fluctuations in data, with the rate of hyperopia being 20.93% in our entire cohort, 60.0% in our Hispanic population, and 9.4% among our non-Hispanic population. Depending on which race or ethnicity was being considered, hyperopia prevalence in our patients can be interpreted as higher or lower than what is reported in the literature.

The function of *ANKRD11* when it comes to development of the central nervous system is not clear. Studies have shown that *ANKRD11* is expressed in the brain and is present in nuclear inclusions in nonneuronal cells [4,28]. This gene has also shown evidence of playing a role in craniofacial development, as *ANKRD11* increases acetylation of p53, which is involved in the pathogenesis of craniofacial developmental syndromes [29,30]. Both disruptions in the central nervous system and craniofacial development may explain a mechanism by which those with KBG syndrome develop ophthalmological conditions. Other related syndromes, including those with 16q24.3 microdeletion syndrome, have shown to overlap clinically with KBG syndrome while also showing ophthalmological abnormalities, including strabismus and refractive errors [17,31,32].

Regardless, a pattern of visual abnormalities exists within our cohort of 43 patients with KBG syndrome, and further research is indicated to investigate these associations.

## 5 LIMITATIONS

The main limitation of our study is that we were unable to provide an age-matched control group from the same interview group without KBG syndrome, leaving us limited to a descriptive statistical analysis between our prevalence data and previously published studies.

Another limitation is that documentation of ophthalmological conditions was done mostly by self-reporting by the participants themselves. Medical records were collected and most provided documentation of visual impairment, but many did not have records from an optometrist or ophthalmologist showing exactly to what extent their visual impairment deviated (in diopters, for example) from what is considered healthy vision. Furthermore, proband 24 provided medical documents showing that there was a mutation in *ANKRD11*, but the exact cDNA and protein change is unknown at this time.

A final limitation lies in the method by which we defined and compared refractive errors and astigmatism. Since inclusion of these conditions were given after a self-report by the interviewed patient, and since most of them had no documentation to provide the extent of refractive error (in diopters) or astigmatism (in spherical equivalents), it was hard for us to have the same inclusion criteria as the studies we compared our prevalence data to. The studies used had different thresholds for which a patient was determined to have a refractive error worth reporting. For example, one study defined astigmatism as a cylindrical refractive error equal to or greater than 1.50 diopters (D) in the worse eye, producing a prevalence of 16.8% among Hispanic children [24]. However, the prevalence of astigmatism with a worse eye cylindrical refractive error equal to or greater than 3.00 D in Hispanic children was determined to be 2.9% [24]. Without our own refractive measurements of our cohort to define the threshold of significant refractive error, and the fact that studies used different thresholds to determine significance, we were left limited in our comparisons of refractive error.

## 6 CONCLUSIONS

While the presentation of KBG syndrome is classically associated with dental, developmental, skeletal, and craniofacial abnormalities, this analysis demonstrates that in a cohort of 43 individuals with diagnosed KBG syndrome, the occurrence of ophthalmological conditions occurs at a rate that is higher than other studied populations. Specifically, the prevalence of strabismus, astigmatism, and myopia among our patient population was higher than what was reported in other studies. The prevalence of hyperopia can be interpreted as higher or lower than what is reported in the literature depending on ethnicity. Nonetheless, more research is needed to determine what role *ANKRD11* mutations have in the clinical manifestation of KBG syndrome, and if altered *ANKRD11* function leads to impaired development of the visual system. If such a connection exists between KBG syndrome and ophthalmological conditions, patients with diagnosed KBG syndrome should receive prompt referral to ophthalmology for evaluation of misalignment or refractive error. Prompt intervention in pediatric patients with misalignment can prevent permanent issues and intervention in refractive errors in patients of all ages can improve their quality of life and function.

## Data Availability

All data produced in the present study are available upon reasonable request to the authors

## AUTHOR CONTRIBUTIONS

GJL conducted all virtual interviews with participants and was responsible for primary data collection. EM was responsible for summarizing primary data. DC was responsible for data analysis and project conception along with GJL. The first draft of the manuscript was written by DC, with critical revision performed by OK, KS, and GJL at several points.

## ACKNOWLEDGMENTS

We would like to thank all families who participated in this study, as well as the KBG foundation for referrals.

## ETHICAL APPROVAL

Both oral and written patient consent were obtained for research and publication, with approval of protocol #7659 for the Jervis Clinic by the New York State Psychiatric Institute - Columbia University Department of Psychiatry Institutional Review Board.

